# Computational analysis of pathological image enables interpretable prediction for microsatellite instability

**DOI:** 10.1101/2020.12.07.20244616

**Authors:** Jin Zhu, Wangwei Wu, Yuting Zhang, Shiyun Lin, Yukang Jiang, Ruixian Liu, Xueqin Wang, Heping Zhang

## Abstract

**Objective:** Microsatellite instability (MSI) is associated with several tumor types and its status has become increasingly vital in guiding patient treatment decisions. However, in clinical practice, distinguishing MSI from its counterpart is challenging since the diagnosis of MSI requires additional genetic or immunohistochemical tests. In this study, we aimed to establishe an interpretable pathological image analysis strategies to help medical experts to identify MSI automatically.

**Design:** Three cohorts of Haematoxylin and eosin-stained whole-slide images from 1033 patients with different tumor types were collected from The Cancer Genome Atlas. These images were preprocessed and tessallated into small tiles. A image-level interpretable deep learning model and a feature-level interpretable random forest model were built up on these files.

**Results:** Both models performed well in the three datasets and achieved image-level and feature-level interpretability repectively. Importantly, both from the image-level and feature-level interpretability, color features and texture characteristics are shown to contribute the most to the MSI prediction. Based on them, we established an interpretable classification framework. Therefore, the classification models under the proposed framework can serve as an efficient tool for predicting the MSI status of patients.

**Conclusion:** This study establishes a interpretable classification framework to for predicting the MSI status of patients and provide more insights to pathologists with clinical understanding.

## Introduction

Microsatellite instability (MSI) is the condition of genetic hypermutability that results from impaired DNA mismatch repair. Cells with abnormally functioning mismatch repair are unable to correct errors that occur during DNA replication and consequently accumulate errors. MSI has been frequently observed within several types of cancer, most commonly in colorectal, endometrial, and gastric adenocarcinomas (1). The clinical significance of MSI has been well described in colorectal cancer (CC), as patients with MSI-high colorectal tumors have been shown to have improved prognosis compared with those with MSS (microsatellite stable) tumors (2). In 2017, the U.S. Food and Drug Administration approved anti-programmed cell death-1 immunotherapy for mismatch repair deficiency/MSI-high refractory or metastatic solid tumors, making the evaluation of DNA mismatch repair deficiency an important clinical task. However, in clinical practice, not every patient is tested for MSI, because this requires additional next-generation sequencing (3, 4), polymerase chain reaction (5) or immunohistochemical tests (1, 6, 7). Thus, it is in high demand for a cheap, effective, and convenient classifier to assist experts in distinguishing MSI vs MSS.

Numerous publications have identified histologic features which are more commonly seen in MSI. By far, it is a well-known fact that tumors have undifferentiated morphology, poor differentiation and the high infiltration of TIL cells are more likely to be MSI (8-11). Unfortunately, it is still challenging to distinguish MSS from MSI based on pathologist’s visual inspections from pathological images since the morphology of MSS is similar to that of MSI (12). The recent technical development of high-throughput whole-slide scanners has enabled effective and fast digitalization of histological slides to generate whole-slide images (WSI). More importantly, the thriving of various machine learning (ML) methods in image processing, makes this task accessible. In recent years, ML has been broadly deployed as a diagnostic tool in pathology (13, 14). For example, Osamu Iizuka et al. built up convolutional neural networks (CNNs) and recurrent neural networks to classify WSI into adenocarcinoma, adenoma, and non-neoplastic (15). The study by Yaniv Bar et al. demonstrated the efficacy of the computational pathology framework in the non-medical image databases by training a model in chest pathology identification (16). Notably, (17, 18) showed that deep learning model can predict MSI directly from Haematoxylin and eosin (H&E) histology and reported the network achieved desirable performance in both gastric stomach adenocarcinoma (STAD) and CC (17). These studies attest to the great potential of ML methods in medical research and clinical practice.

There is no doubt that the ML revolution has begun, but the lack of the ‘interpretability’ of ML is of particular concern in healthcare (19, 20). Here, the ‘interpretability’ means that clinical experts and researchers can understand the logic of decision or prediction produced by ML methods (21). In essence, it urges ML systems to follow a fundamental tenet of medical ethics, that is, the disclosure of necessary yet meaningful details about medical treatment to patients (22). Also importantly, interpretability helps clinician understand that the model’s decision would have a potential fairness/bias issue because the samples used in training models are not necessarily representative of the underlying study population (23). In addition, for both scientific reproducibility and medical safety reasons, interpretability allows researchers to know to the extent to which small systematic perturbations can alter the predictions to the input data, which might be generated by measurement biases. Finally, clinical experts are accessible to potentially crucial domain-knowledge hidden in the interpretable ML models (21). Unfortunately to the best of our knowledge, most of the existing MSI diagnosis systems, especially deep-learning based systems, are non-interpretable. Therefore, there is an urgent need to establish a new research paradigm in applying an interpretable ML system in medical pathology field (24-28).

In this study, we used H&E-stained WSI from The Cancer Genome Atlas (TCGA): 360 formalin-fixed paraffin-embedded (FFPE) samples of CC (TCGA-CC-DX) (29), 285 FFPE samples of STAD (TCGA-STAD) (30) and 385 snap-frozen samples of CC (TCGA-CC-KR). H&E stained images in these databases have already been tessellated into 108020 (TCGA-STAD), 139147 (TCGA-CC-KR), and 182403 (TCGA-CC-DX) color-normalized tiles, (17) and all of them only target region with tumor tissue. The aims of the study are: (i) to build an image-based ML method on MSI classification and post-process the fed image to a heat map to interpret the diagnosis of MSI at an image level; (ii) to design a fully transparent feature extraction pipeline and understand the pathological features’ importance and interactions for predicting MSI by training a feature-based ML model.

## Results

### A Deep Learning Classifier and Image-Level Visual Interpretability

We used a commonly used end-to-end CNN, Resnet18 (31). To fit this deep learning (DL) model for different cancer subtypes, we trained three Resnet18 networks based on 70% tiles randomly sampled from three datasets. The remaining 30% tiles in each dataset were used for testing. At the testing time, a patient’s slide was predicted to be MSI if at least half of the tiles were predicted to be MSI. The patient-level accuracy and area under the curve (AUC) were 0.84 in the KR cohort, 0.81 in the DX cohort, and 0.80 in the STAD cohort (Fig. 1b), respectively, in the test set.

**Fig. 1.**
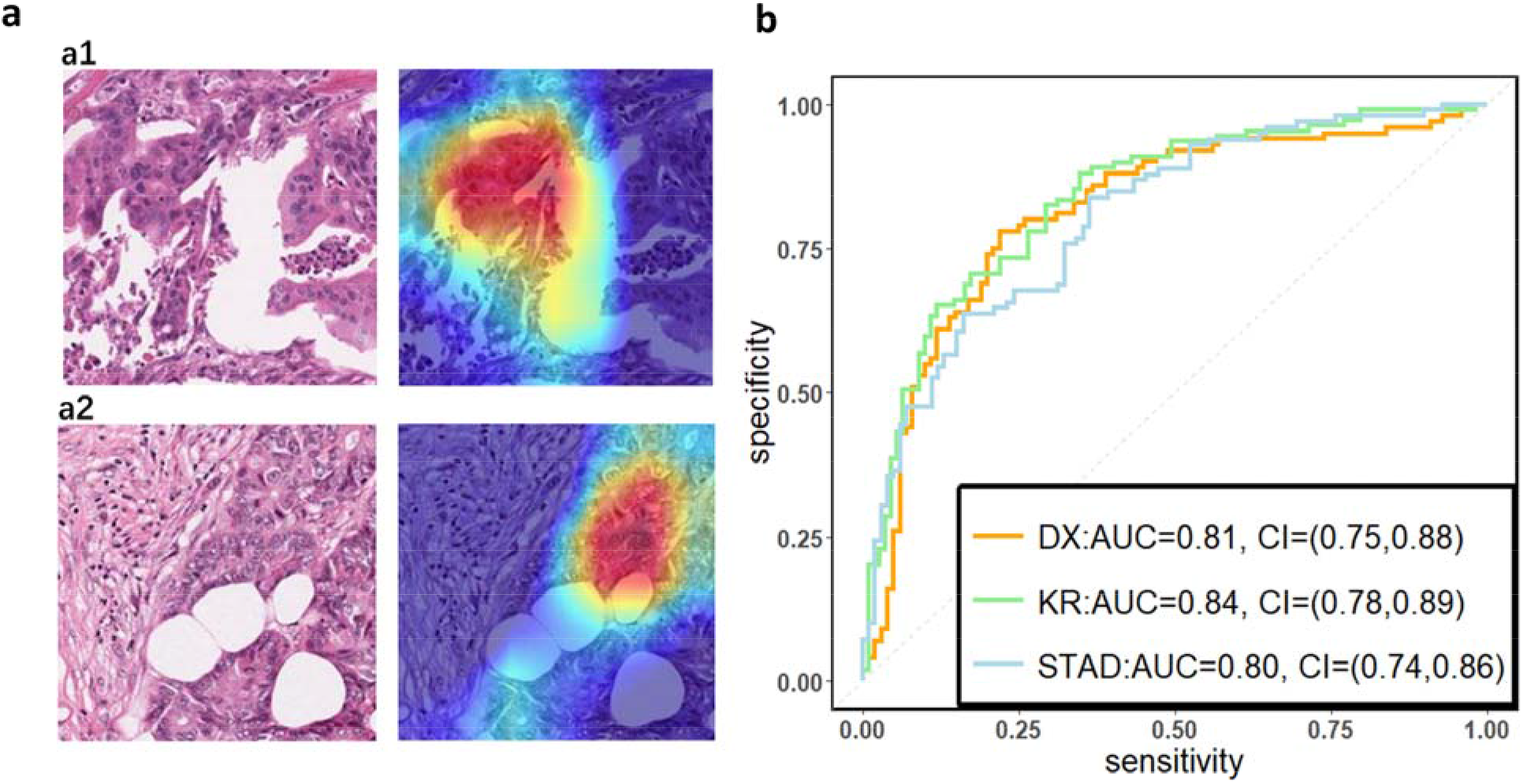
(a) The original tile and the corresponding heatmap output by the GCAM. **(a1)** and **(a2)** display tiles from the TCGA-CC-DX dataset labeled with MSI and MSS, respectively. In the heatmaps, the brighter region contributes more to the classification. For instance, the red one is the most highlighted area, while the blue regions contribute limitedly. **(b) Patient-level receiver operating characteristic (ROC) curve for classifying MSI versus MSS in the three datasets with deep learning**. The 95% confidence intervals (CI) were computed by the bootstrap method.

Based on the trained DL model, the Gradient-weighted Class Activation Mapping (Grad-CAM) was used to make the convolutional based model more transparent by generating localization maps of the important regions (32). To unveil the hidden logic behind the DL and provide visual interpretability, we deployed Grad-CAM to find out which part of the H&E image supports DL’s classification. Two typical images for interpreting DL prediction logic are shown (Fig. 1.a). Our pathologist noted that the highlighted region in Fig.1.a tended to be where immune cells are mainly concentrated in the tumor organism; meanwhile, we also found that the highlighted region presented distinct color and texture characteristics. We were intrigued by this phenomenon and further examined this important region in great detail.

### Transparent Pathological Image Analysis Workflow and Feature-Based Classification Model

The results from Grad-CAM suggested that certain features of the H&E stained images might encode essential regions of the tumor organism. To further investigate this, we developed a multi-step, automatic and transparent workflow (Fig. 2.a). In the first step, we standardized the three image datasets by standard image processing techniques (e.g., white balance and brightness adjustments). After the image pre-processing, we extracted visible pathological features. Motivated by the feedback from Grad-CAM and existing studies, we focused on these H&E feature characteristics: global and local color features in RGB and HSV channels, the numbers of infiltrating immune cells and tumor cells, the grading of differentiation and the texture features from tumor cells. A total of 182 features were extracted from each image tile, and some representative ones are displayed in Fig. 2.b.

**Fig. 2.**
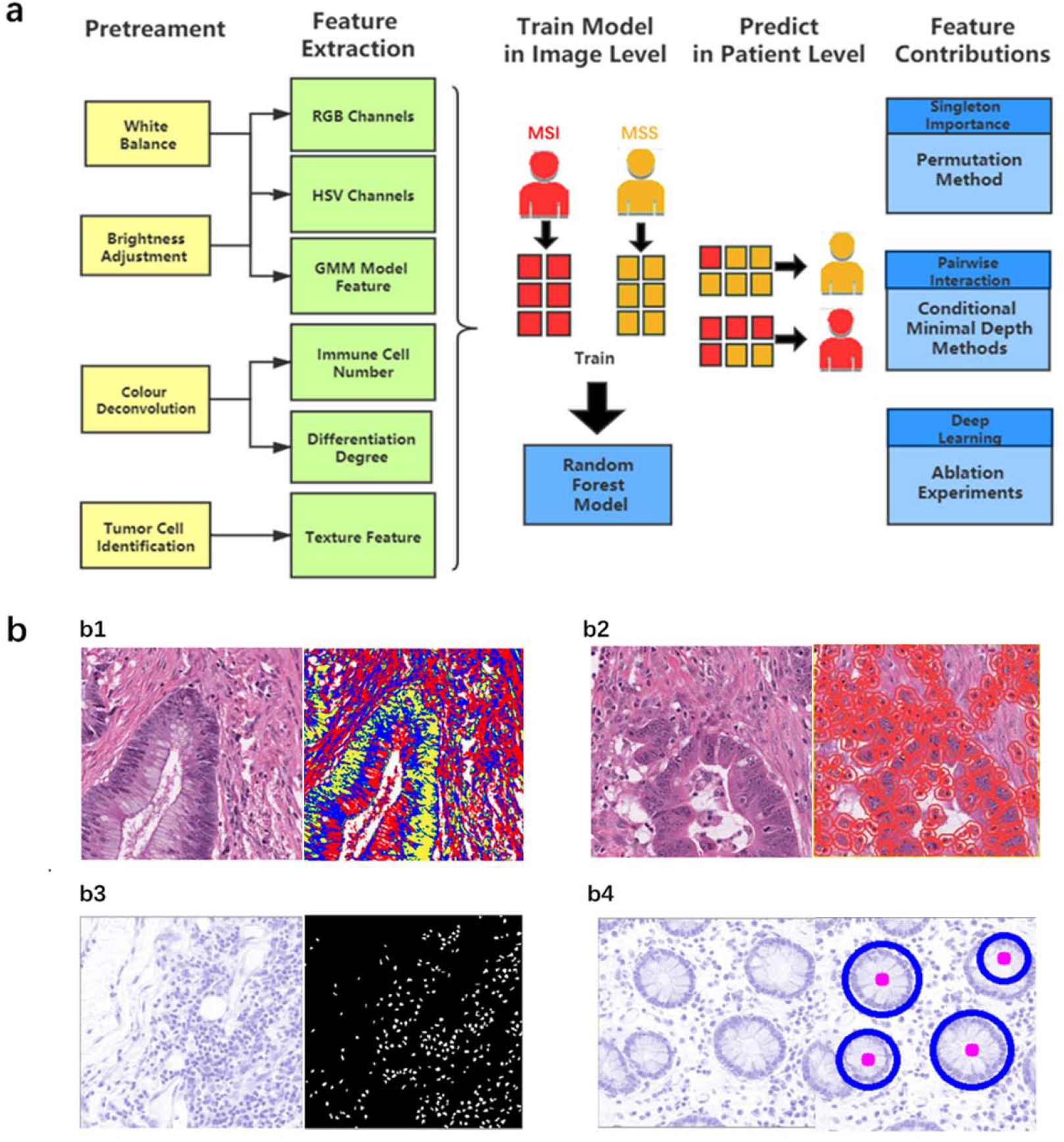
(a) The workflow of studying pathological features in discriminating against MSI from MSS. Five main steps - pretreatments, feature extraction, model training, patient-level predictions, and feature contributions analysis - were sequentially executed to improve image quality, generate pathological features, build statistical model, evaluate model performance, and measure features’ contributions, respectively. **(b) Typical feature extraction result. (b1) GMM model for image segmentation**. The figure on the left is a tile from the TCGA-CC-DX dataset, and its image-segmentation tiles processed by the GMM method are shown in the figure on the right. The green part whose grayscale is the lowest among the three parts tends to be tumor tissue, while the blue and red ones represent non-tumor tissue. **(b2) Tumor cell detection before Haralick texture identification**. The figure on the left is an original tile, while the one on the right is processed with tumor identification. Each red circle in the tile on the right indicates the boundary of one tumor cell. **(b3) Infiltrating immune cells detection**. The detection of immune cells allows us to calculate the connectivity domain. **(b4) The grading of differentiation**. Detect the circularly similar arrangement in one slice and grade the degree of differentiation based on its amount.

We then applied random forest (RF) (33), one of the most popular ML algorithms, to all three databases to classify MSI versus MSS on H&E stained histology slides. We randomly selected 70% of patients in every dataset during training, and all their tiles were used in training while the rest tiles were held out and used as test sets. In the test sets of each dataset, true MSS image tiles cohort had a median MSS score (the proportion of the prediction result judged to be MSS in each decision tree of the forest) that was significantly different from those of MSI tiles (the P-values of the two-tailed test were 0.02, 0.0024, 0.002 in the three datasets), indicating that our models can distinguish MSI from MSS. Since one patient may have many different tiles, we obtained the patient-level MSI scores by averaging the RF’s prediction on all its tiles. AUCs for MSI detection were 0.78 (95% CI 0.7-0.82) in KR cohort, 0.7 (95% CI 0.65-0.74) in DX cohort, and 0.74 (95% CI 0.65-0.79) in STAD cohort (See Fig. 3.b, Fig. S1.b, and Fig. S2.b). These results show that visible pathological features can be useful in MSI prediction.

**Fig. 3.**
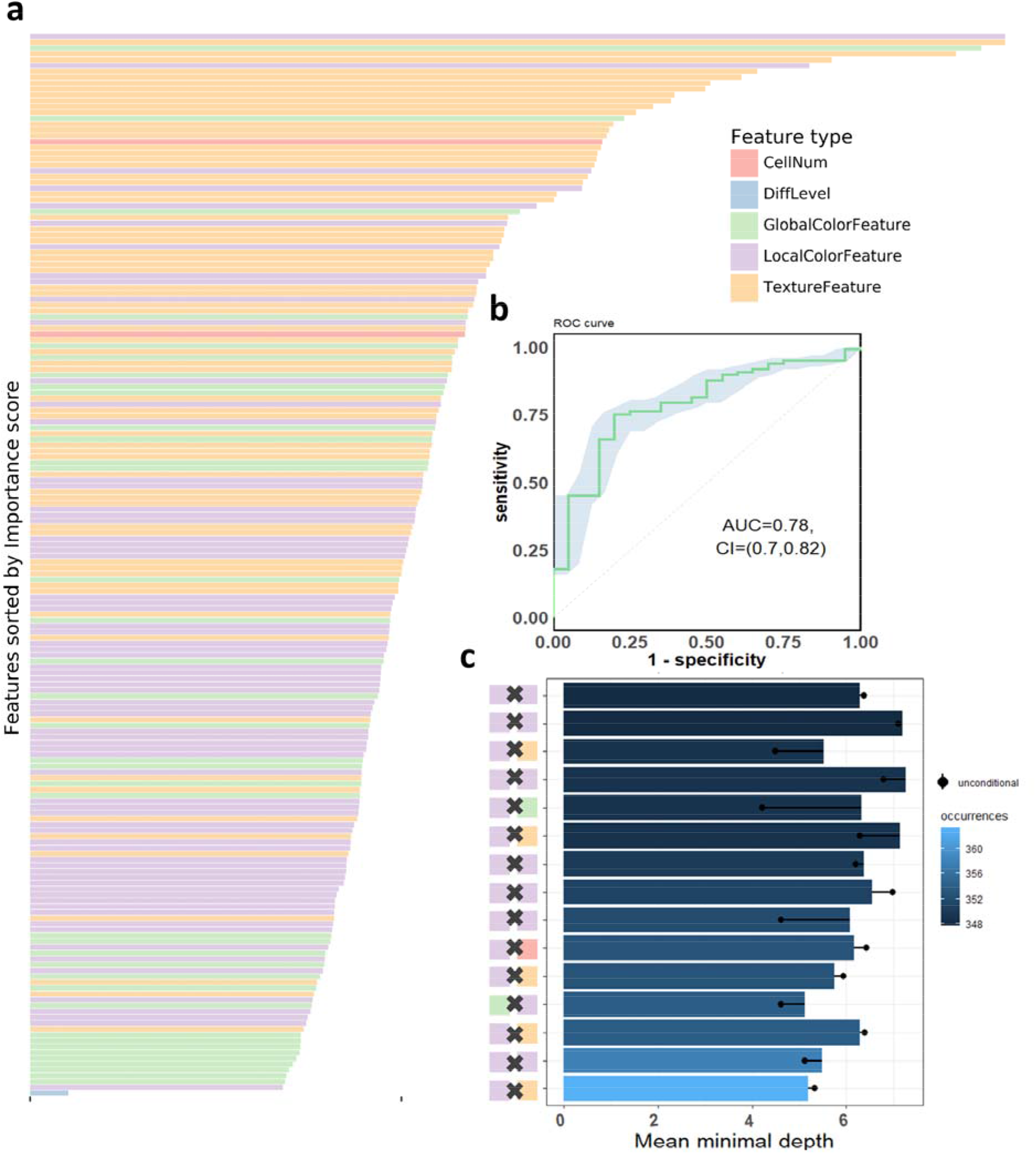
The visualization of performance and interpretability of the RF in KR dataset. **(a) The bar plot of permutation-based variable importance**. Features are arranged from top to bottom in order of importance (the names of the features are provided in the order in Table S2). **(b) The patient-level ROC curve for classifying MSI versus MSS with random forest**. The blue bands are 95% confidence interval (CI) computed by the bootstrap method. **(c) The bar plot of the mean of conditional minimal depth (the top 15 feature pairs of interaction are shown)**. A feature pair of interaction is listed as *A* × *B*, where *A* and *B* are one of feature type and their concrete names are listed in Table S3. Feature pairs are arranged from the bottom to top in the order of the occurrences, which are represented by the color intensity of the bars. The bar’s length indicates the mean of conditional minimal depth and the distance from the dot to the y-axis measures the mean of minimal depth of *B*. The length of the dot line implies the gap between them, measuring the effect of pairwise feature interaction. A large gap implies a strong interaction.

### Feature-Level Visual Interpretability: Feature Importance and Interactions

One of the attractive advantages of RF is that it can evaluate the importance of the features. Therefore, we verify and quantify these features’ power in distinguishing MSI from MSS by extracting information from a trained model. A representative pattern can be discovered from the visualization of permutation-based feature importance (33, 34) in the KR dataset (Fig. 3). From the figure, we can deduce that the texture features play a dominant role. Since the texture features reflect the surface’s average smoothness of the tumor cells in one tile, we deduce that the characteristics of the tumor surface are an important clinical indicator in automatic MSI diagnosis. Color features also have important contributions. In the global color feature, the higher-order statistics (skew and kurtosis) contribute more than the first-order statistics (mean and quantile), indicating that some useful information contributing to classification are hidden in high order features. Local color features also deserve our attention. Compared to global color features, the local ones were useful in image segmentation by dividing slices into different clusters, and we obtained the information in each cluster. Fig. 2.b demonstrates the clinical utility of the clusters as they closely reflected tumor tissue versus non-tumor tissue. The number of infiltrating immune cells was also important as expected, whereas the differentiation grade contributed the least in every dataset.

It is widely accepted that feature interactions (i.e., the joint effect of features) can be important for the complex disease (35-38). Our feature-based RF models also allow us to exploit the pairwise feature interactions in MSI classification, and thus, we can attain a more clear understanding of the characteristics of MSI tiles and the mechanism of RF. Here, we use conditional minimal depth (39) to quantitatively assess feature interaction and then demonstrate the foremost 15 pairwise interactions (Fig. 3.c, Fig. S1.c, Fig. S2.c). The feature type with the most effective interaction effect with other features in each dataset are: the local color feature in KR, the global color feature in DX, and texture features in STAD. The three features enhanced the importance of the features interacting with them, even the features themselves may have a weak effect before. It is also worthy to note that interactions incline to occur more often between color features and texture features or between local color and global color features. To understand how the paired features jointly help the MSI diagnosis, we plot the prediction values of typical feature interaction on a grid diagram (Fig. S3). In the KR dataset, a greater immune cell number and a lower value of the 75th percentile of red channel lead to a higher probability of MSS. In DX, a higher value of the max caliper in tumor cells and a fewer tumor cell number lead to a higher probability of MSS. In STAD, a lower value of the optical density range of tumor cells’ nucleus in Hematoxylin stains and a higher value of texture feature correlation in eosin stains lead to a higher probability of MSS.

## Discussion

To our knowledge, this is the first study to not only build up a classification model in distinguishing MSI from MSS but also provide a detailed interpretability analysis. Previous studies in investigating the pathologic predictors of microsatellite instability through feature extraction and logistics regression model suffered from the limited learning capability as well as the small sample size, and thus could not achieve satisfactory performance (9). Other works on MSI classification paid attention to the enhancement of the prediction accuracy by establishing a DL network but did not provide a detailed description of the mechanism behind the model (17). In this study, we tackled these problems through using three different cancer types datasets from TCGA and following the framework of interpretability with two steps: first, built up a high-performance DL network with a visual explanation capacity as model-based interpretability; secondly, we further analyzed and confirmed features’ power using a feature-based interpretable model.

To build an interpretable DL network, we trained residual learning CNNs and deployed Grad-CAM to the final convolutional layer of the network to produce the heatmap that reflects the highly-contributed region. Notably, through its coarse localization map of the image’s essential regions, it provided preliminary insight into highly-contributed pathological features. To understand the contribution of the pathological features on MSI classification, we extracted the clinically meaningful features, trained an RF classifier based on those features, and assessed the importance of those features, and exploited their interaction. Notably, we found that the texture and color of the H&E image and the interactions among them were crucial for diagnosing MSI. To the best of our knowledge, this has not been noted before. Another interesting fact is that, the feature type tend to interact with the other features has a clear difference in the three datasets due to the image heterogeneity raised from the diversity of cancer type (CC or STAD) and tissue preservation methods (snap-frozen or FFPE) (40), indicating that the feature interaction mode was influenced by preservation methods and tumor types. Yet this insight would not be attained from “black-box” machine learning method. Moreover, we hypothesized that the dominant-role features such as color in RF models were also important in the DL model. To test our hypothesis, we eliminated the mean color differences between MSI and MSS groups and reevaluated our DL models’ AUCs. Specifically, we calculated the RGB mean value of all tiles in both groups and centralized the RGB mean value of every tile into that population mean value. We found that the AUCs were reduced by 0.11, 0.12, and 0.14 in DX, KR, and STAD datasets, respectively, supporting our hypothesis that color features also contributed to the DL model.

We note that our findings warrant replications through further biological experiments. The H&E stain is capable of highlighting the fine structures of cells and tissues. Most cellular organelles and extracellular matrix are eosinophilic, while the nucleus, rough endoplasmic reticulum, and ribosomes are basophilic. Our study shows that the spectrum, intensity, and texture of colors matter in distinguishing MSI from MSS, which needs further validation. We hypothesize that MSI tumor usually has distinct color/texture characteristics due to diverse gene mutation pattern (41, 42). Another limitation of this study is that the cases in TCGA datasets may not be an unbiased collection from the real situation since pathologists may only upload the representative ones. Although our model performed well in these histopathology images, we should admit that their performance in the actual clinical settings requires further research. Another limitation is that our study only focused on H&E stained images, and we could not confirm whether the pattern in this study, especially the color features’ contribution, works in other types of histopathology slices. The classifier model, under other types i.e., immunochemical stained images, remains to be explored and established.

Further, our framework provides a positive feedback cycle in assisting pathologist’s diagnosis of MSI (Fig. 4). Specifically, the localization map outputted by our DL models can help experts to narrow their focus on the specific region of the whole H&E slide, thereby contributing to a more accurate and apprehensible diagnosis with the prediction result of our model. The features’ distribution under our interpretable model can provide experts with more insight into analyzing the slices of MSI and MSS from clinical perspectives. Further, considering the similar feature distribution pattern in three datasets we used, it is possible that after running the same pipeline on MSI H&E slides under different cancer types, we can discover a generalization pattern behind them. After training on a larger dataset, the accuracy of the identification and the interpretability could improve, thereby contributing to accurate sample curation and treatment development of this aggressive cancer subtype.

**Fig. 4.**
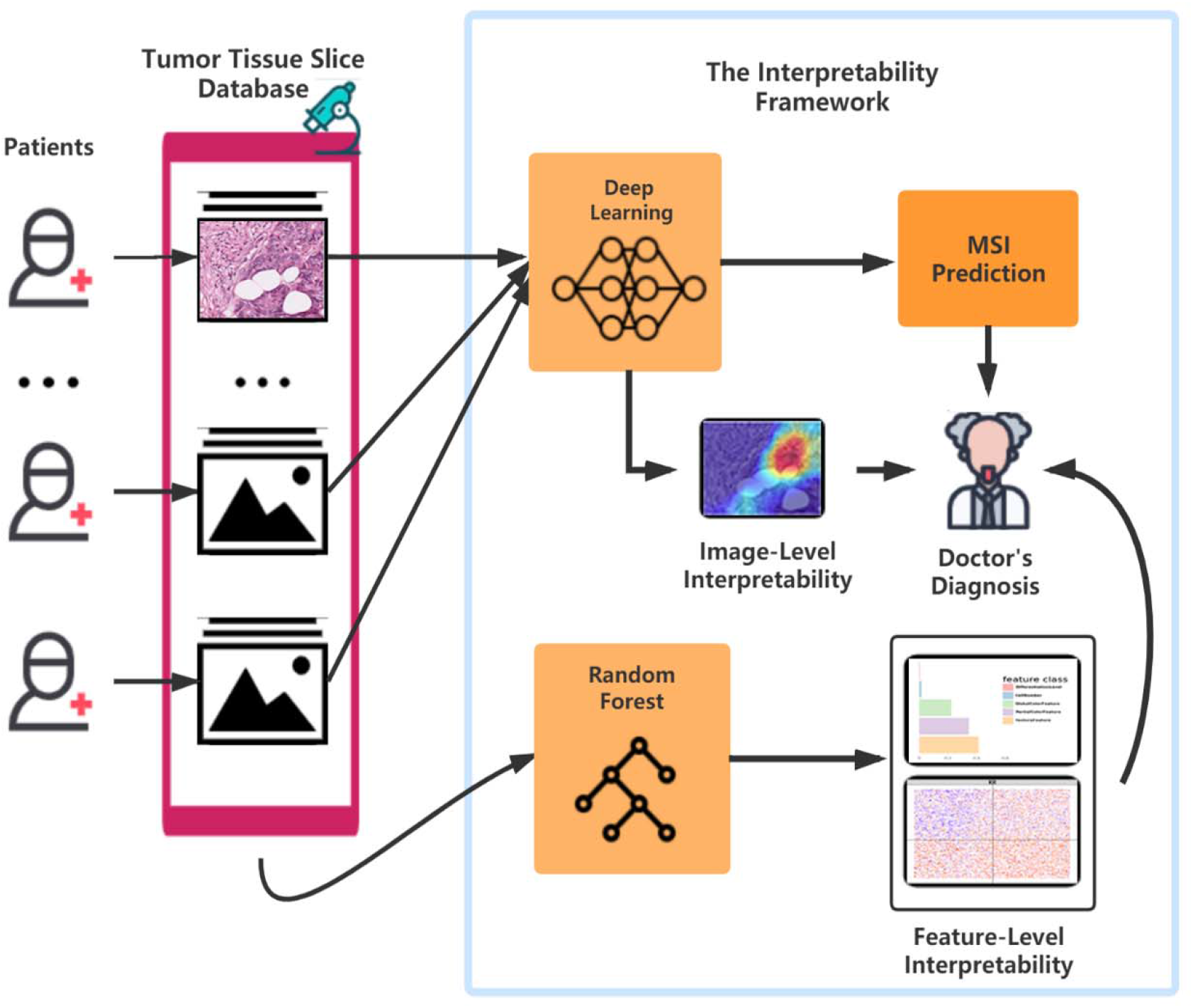
The flowchart of the pattern in which our framework can assist the doctor’s diagnosis. After surgery or biopsy, the embed cut H&E provided by each patient would go through MSI screening with deep learning. The doctor can make a critical diagnosis based on his insight combined with the prediction result and the deep learning model’s visualization. Meanwhile, with the amplifying of the H&E datasets, the random forest could develop a more precise and interpretable model, which helps the doctors detect MSI.

In summary, we developed ML models with decent power in the prediction of MSI. Moreover, our models exhibit a visual heatmap demonstrating high-contribution regions for MSI prediction in the H&E image. We certified certain pathological features with non-trivial importance in MSI classification, which is not explicitly studied in the previous research. Therefore, our study facilitates MSI diagnosis based on H&E image and sheds light on the understanding of MSI at both image-level and features level. As a by-production of our study, a user-friendly and ongoing-upgraded web application (http://14.215.135.56:3838/DL/) was developed for world-wide clinical researchers.

## Materials and Methods

### Histopathology Image Sources

The whole-slide H&E stained histopathology images were obtained from TCGA, including three cancer subtype datasets. Dataset DX consisted of 295 MSS patients and 65 MSI patients from FFPE samples of CC. Dataset KR contained 316 MSS patients and 72 MSI patients from snap-frozen samples of CC. Dataset STAD collected 225 MSS patients and 60 MSI patients of FFPE stomach adenocarcinoma.

All the images used in our models have already gone through tumor tissue detection and have been tessellated into small tiles in J.N. Kather’s work (https://zenodo.org/record/2530835 and https://doi.org/10.5281/zenodo.2532612). There are 108020 tiles in TCGA-STAD cohort, 139147 in TCGA-CC-KR, and 182403 in TCGA-CC-DX. Color normalization has already been performed on every tile using the Macenko method (43), which converts all images to a reference color space.

### Details of DL and Grad-CAM

We built the Resnet18 in Python 3.7 with TensorFlow-GPU 1.14.0 and Keras 2.3.0. The patient-level AUCs, ROC curves, and 95% stratified bootstrap CIs for ROC curves were computed and visualized by two R packages: pROC (44) and ggplot2 (45). Grad-CAM utilizes the gradient information abundant in the last convolutional layer of a CNN and generates a rough localization map of the important regions in the image. We apply the rectified linear unit to the linear combination of maps to generate localization maps of the desired class. Grad-CAM visualization was implemented in Python 3.7 with TensorFlow-GPU 1.14.0 and Keras 2.3.0.

### Image Pretreatment

We apply pretreatments to the tiles before feature extraction. First, in order to avoid the influence of color cast, the natural appearance tone of the object is altered in the formation of images when exposed in a lightning condition of different color temperature, white balance is performed on our cohorts. Since every tile has an area without cell organization, i.e., without H&E stained, we could view that part as the neutral reference in adjustment. Besides the color cast, overexposure and underexposure also may result in the distortion of our features. Still, taking the unstained area as the reference, we regulated all tiles into the same level of brightness. In addition, to get the location of immune cells’ nuclei, we need to perform color deconvolution (46), an algorithm used to separate color space from immunohistochemical staining on every tile of our datasets. To realize it, software ImageJ (47) was used with a plugin called Color Deconvolution. Besides, to extract the Haralick texture features (48) of tumor cells, we used a positive cell detection plugin in QuPath software to locate every tumor cell in each tile and use its batch process to get needed features.

### Feature Extraction

In global color feature extraction, the region of interest (ROI) is a stained area. We recorded mean value, quantiles (25%, 50%, 70%), and higher-order moments (variance, kurtosis, and skewness) in ROI of each channel in RGB and HSV as our global features. Besides, with Gaussian mixture model (GMM) model (49), we perform image segmentation to each tile to divide the ROI into three clusters and record the corresponding features in every cluster as our local features. We located immune cells’ nuclei after color deconvolution according to their size and grayscale and calculated the amount as the feature. As for the differentiation degree of tissue in tiles, we performed dilation, erosion, and circle Hough transforms (50) to identify outlines similar to circle in images, to decide their differentiation degree. Since the more regular shapes exist, the more highly the tissue differentiates. Since we have recorded the tumor cell’s location, we extract Haralick features of each tumor cell in one tile and adopt the mean value of all cells’ as this tile feature via QuPath software (51). Besides, we also recorded the count of a tumor cell as our feature. The Wilcoxon rank-sum test is applied to all features, and most of them are significantly different between MSI and MSS in image level (Table S1).

### Random Forest Model for MSS and MSI Classification

Our RF method was built and tested using Python version 3.7.1 with RandomForestClassifier in sklearn.ensemble library (52). During training, 70% of patients in every dataset were randomly selected, and all of their tiles were used in training while the rest tiles were held out and used as test sets. There are some anomalous tiles in each dataset, i.e., blurred or color disorder, resulting in the loss of the information contained in them. Therefore, we disposed of all of them in every dataset. In addition, we also delete the tiles owning an extreme immune cell number (a value that significant in 1% level) since an extremely small number may represent the non-tumor area while a too large number represents lymphatic concentration area. In each forest, we set 500 trees in total and take Gini impurity as the criterion. The minimum number of samples to split an internal node is 2, and other parameters follow the default setting. In all cases, training and test sets were split on a patient level, and no image tiles from test patients were present in any training sets. We used pROC and ggplot2 packages to assess and visualize the model performance. And to further analyze the feature importance and interactions between features, we also used R version 3.5.1 with randomForest package (53) to rebuild that random forest and analyze and visualize the relations between different features with randomForestExplainer package (54).

### Permutation Feature Importance and Contional Minimal Depth

Permutation-based feature importance (33) is a widely-used model inspection technique for RF. It is defined to be the decline in a model accuracy when one feature’s values are randomly shuffled. The shuffle procedure cancels the relationship between the label and the feature, and thus, the drop in the model accuracy can serve as a measurement for the importance of the feature in RF. An alternative feature importance, minimal depth (39), is defined as the depth when a feature splits for the first time in a tree. For example, if a feature splits the root node in a tree, then its minimal depth is 0. The mean of minimal depths over all trees in a forest can measure the feature importance. The importance ordering of features under it keeps highly consistent with the result from the permutation-based method (Fig. S4).

To investigate the interaction between two different features, we used a generalization of minimal depth, conditional minimal depth, that measures the depth of the second feature in a subtree with the root node where the first feature splits (54). Specifically, we recorded all of such splits with the first feature and calculated the mean of conditional minimal depths of the second features given the first feature. A large gap between the mean of conditional minimal depth and the mean of minimal depth implies possibilities for the second feature being used for splitting after the first feature. The occurence of the large gap implies the two features have a strong interaction.

### Ablation Experiment for Deep Learning

We eliminated the RGB mean differences between MSI and MSS groups in the test set by adjusting the mean value in each tile in the test set to the mean value of all the tiles as a whole. Then we feed the adjusted tiles in the test set into the trained neural network. The drops of AUCs after revaluation can verify the contribution of the RGB feature in the classification of the DL network.

## Data Availability

Data, models, or code generated or used during the study are available in a repository in accordance with funder data retention policies.

## Acknowledgments

Wang’s research is partially supported by The National Key Research and Development Program of China (2018YFC1315400), NSFC (11771462, 71991474), The Key Research and Development Program of Guangdong, China (2019B020228001), and the Pearl River S&T Nova Program of Guangzhou (201806010142). Liu’s research is supported by the National Natural Science Foundation of China (81902381). Zhang’s research is supported in part by U.S. National Institutes of Health (R01HG010171 and R01MH116527).

## Notes

### Competing Interest Statement

The authors have declared no competing interest.

### Funding Statement

No external funding was received in this study

### Author Declarations

This research involves analysis of de-identified data initially collected by The Cancer Genome Atlas Program. The Institutional Review Board of Sun Yat-sen University reviewed this research and found that it is exempt since the research does not involve identifiable private information.

