## supplementary1 for "Computational analysis of pathological image enables interpretable prediction for microsatellite instability"

**This PDF file includes:**

Figures S1 to S4

Tables S1 to S4

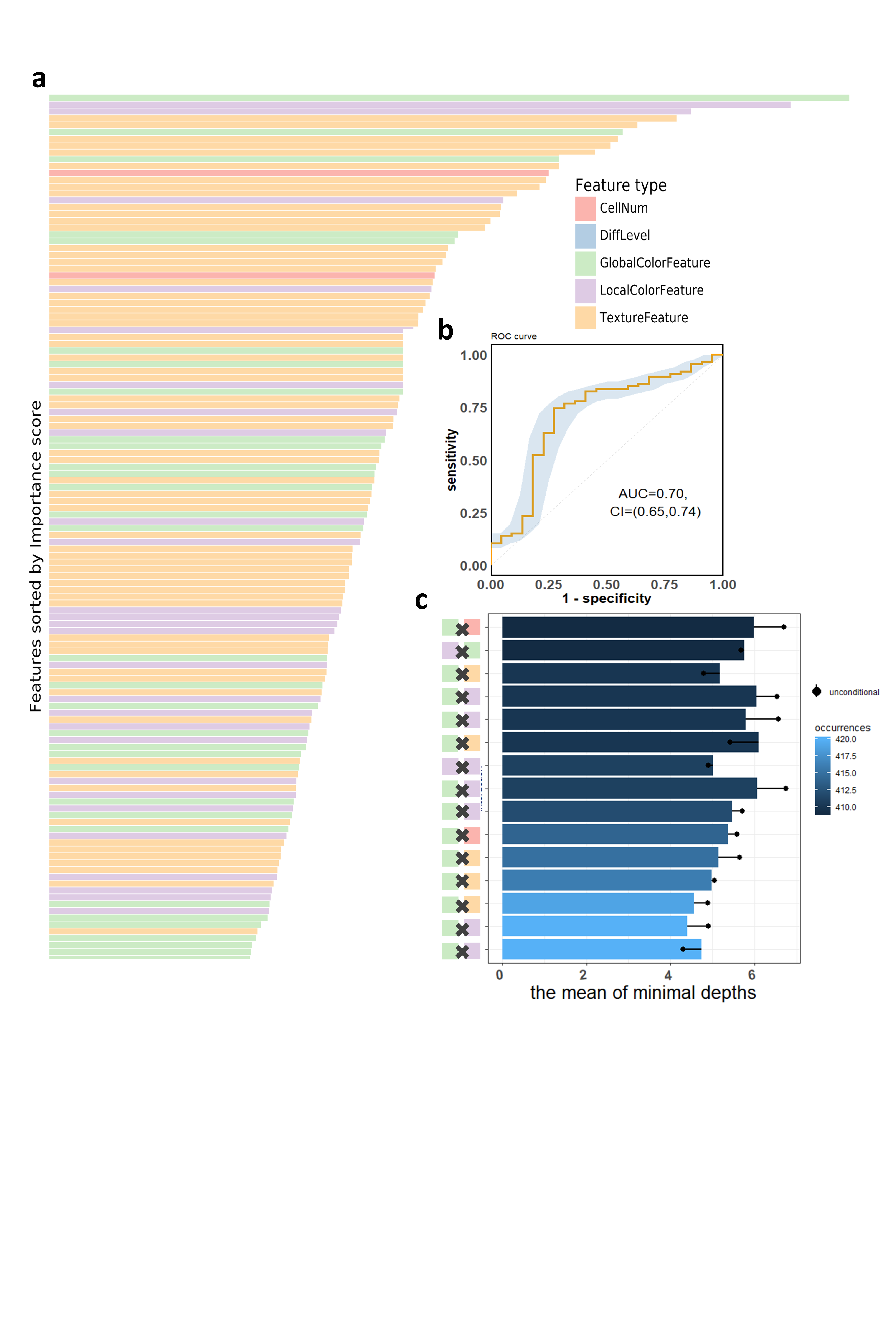
**Fig. S1. The visualization of performance and interpretability of the RF in DX dataset. (a) The bar plot of permutation-based variable importance.** Features are arranged from top to bottom in order of importance (the names of the features are provided in the order in Table S2). **(b) The patient-level ROC curve for classifying MSI versus MSS with random forest.** The blue bands are 95% confidence interval (CI) computed by the bootstrap method. **(c) The bar plot of the mean of conditional minimal depth (the top 15 feature pairs of interaction are shown).** A feature pair of interaction is listed as *A* × *B,* where *A* and *B* are one of feature type and their concrete names are listed in Table S3. Feature pairs are arranged from the bottom to top in the order of the occurrences, which are represented by the color intensity of the bars. The bar's length indicates the mean of conditional minimal depth and the distance from the dot to the y-axis measures the mean of minimal depth of *B*. The length of the dot line implies the gap between them, measuring the effect of pairwise feature interaction. A large gap implies a strong interaction.

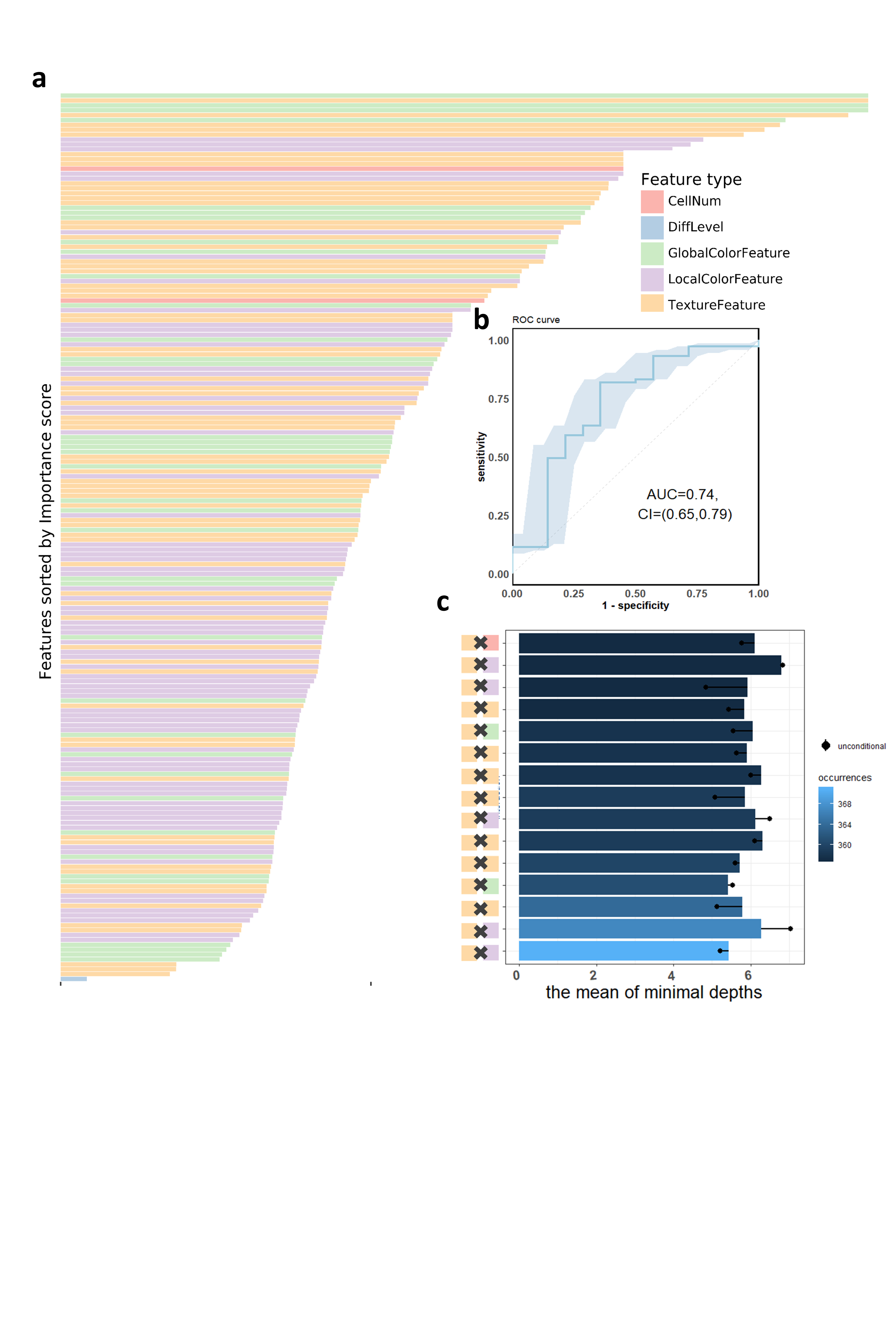

**Fig. S2.** **The visualization of performance and interpretability of the RF in STAD dataset.** **(a) The bar plot of permutation-based variable importance.** Features are arranged from top to bottom in order of importance (the names of the features are provided in the order in Table S2). **(b) The** **patient-level ROC curve for classifying MSI versus MSS with random forest.** The blue bands are 95% confidence interval (CI) computed by the bootstrap method. **(c) The bar plot of the mean of conditional minimal depth (the top 15 feature pairs of interaction are shown).** A feature pair of interaction is listed as *A* × *B,* where *A* and *B* are one of feature type and their concrete names are listed in Table S3. Feature pairs are arranged from the bottom to top in the order of the occurrences, which are represented by the color intensity of the bars. The bar's length indicates the mean of conditional minimal depth and the distance from the dot to the y-axis measures the mean of minimal depth of *B*. The length of the dot line implies the gap between them, measuring the effect of pairwise feature interaction. A large gap implies a strong interaction.

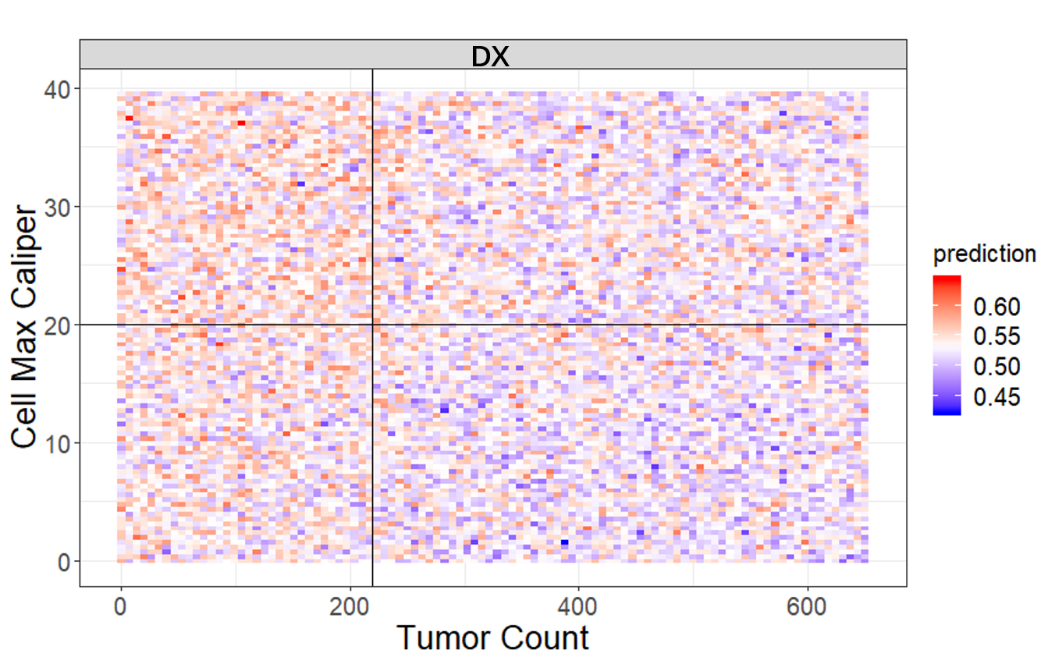

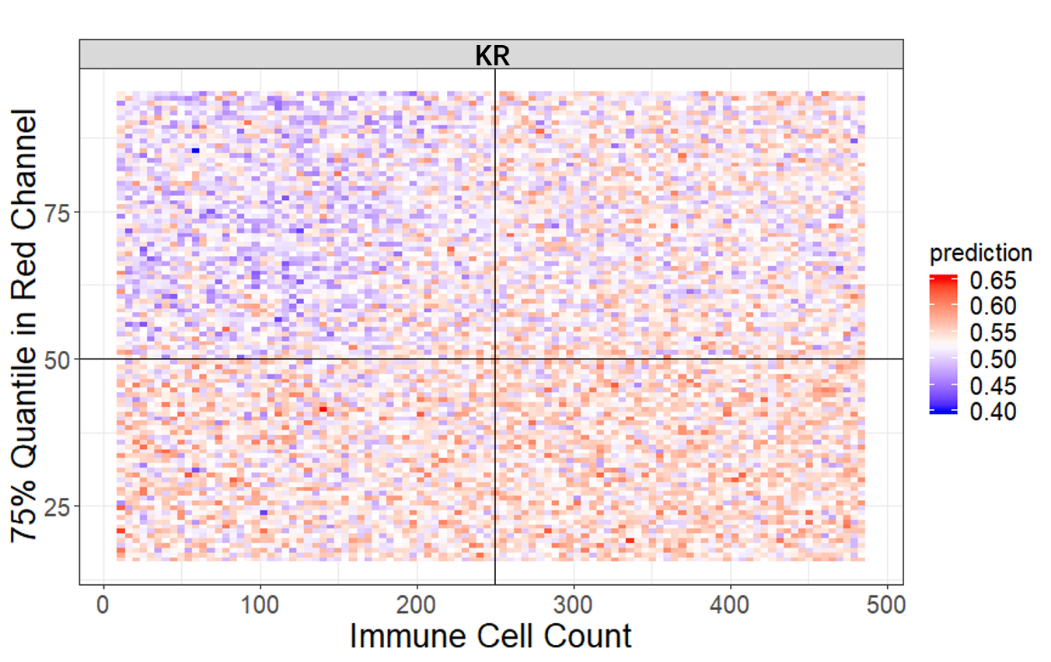

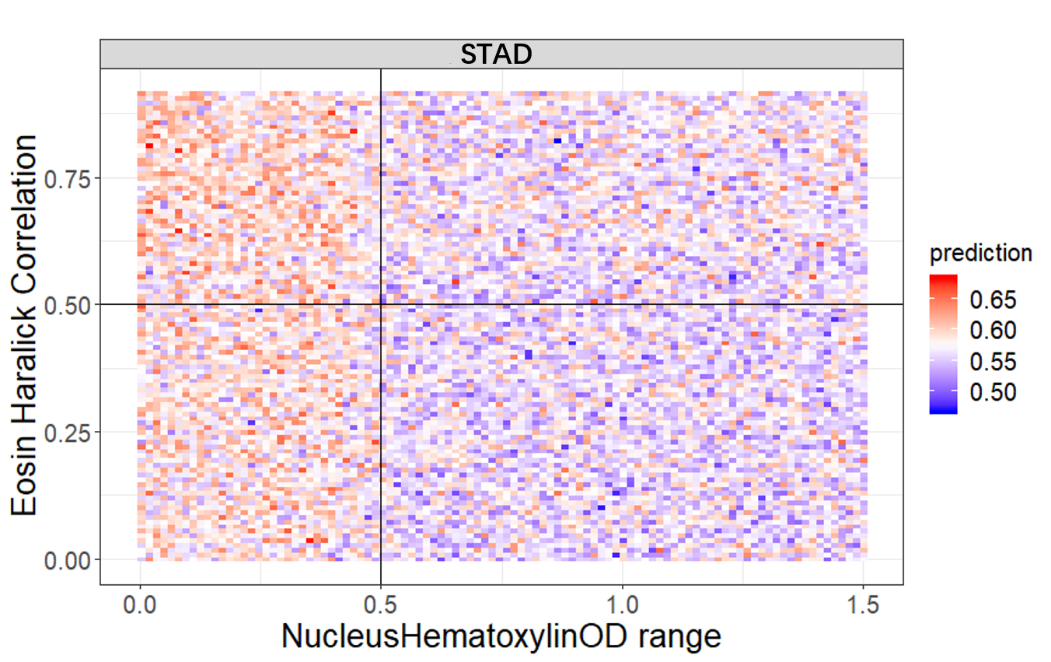

Fig. S3. The visualization of typical pairwise features’ interaction in each dataset. Each panel visualize the random forest’s prediction on a grid of values of a certain interaction. The two axes represent the two corresponding features. The prediction value ranges from 0 to 1 with color from blue to red. The bluer means a larger probability of MSI while the redder tends to be MSS.

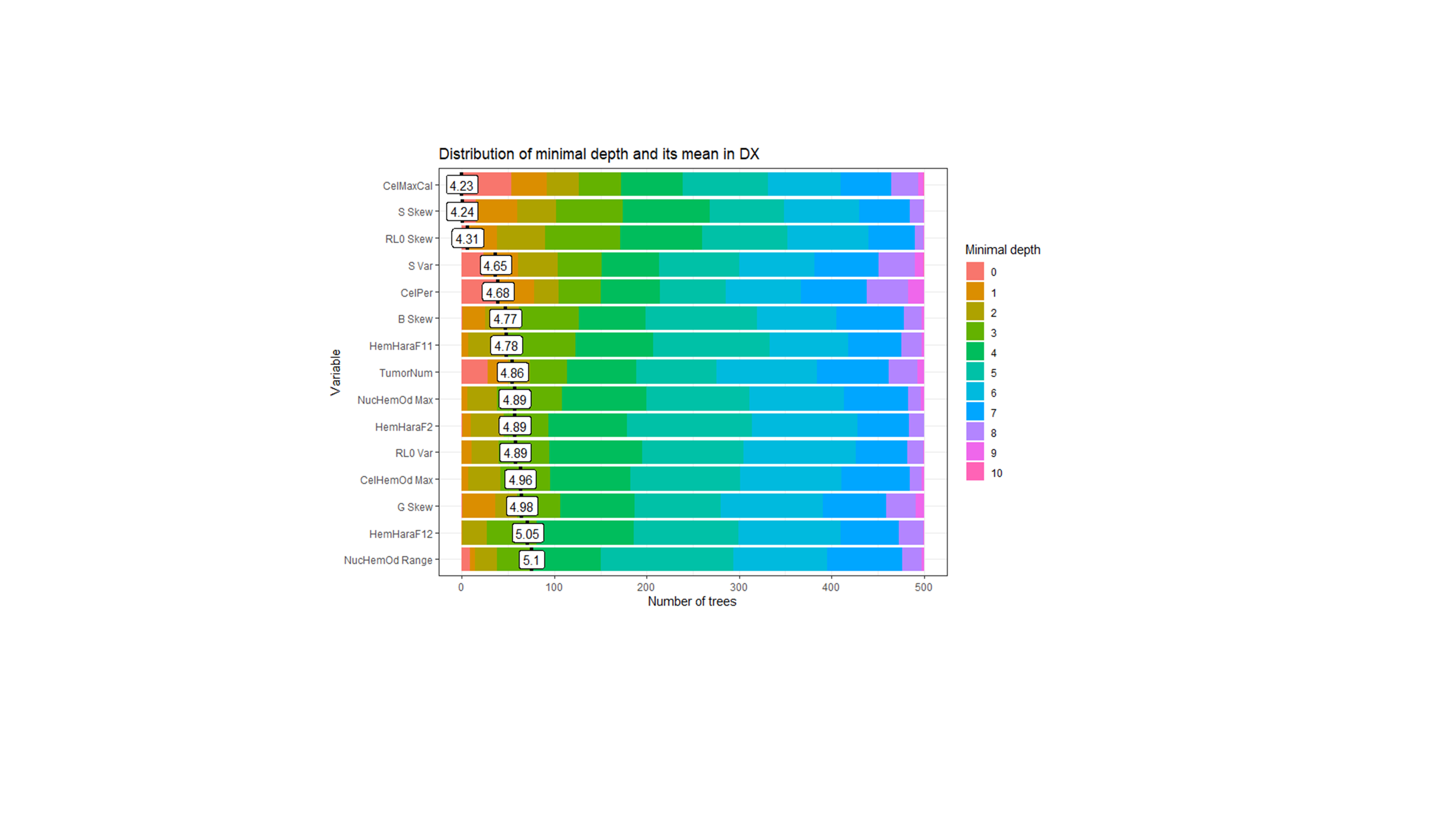

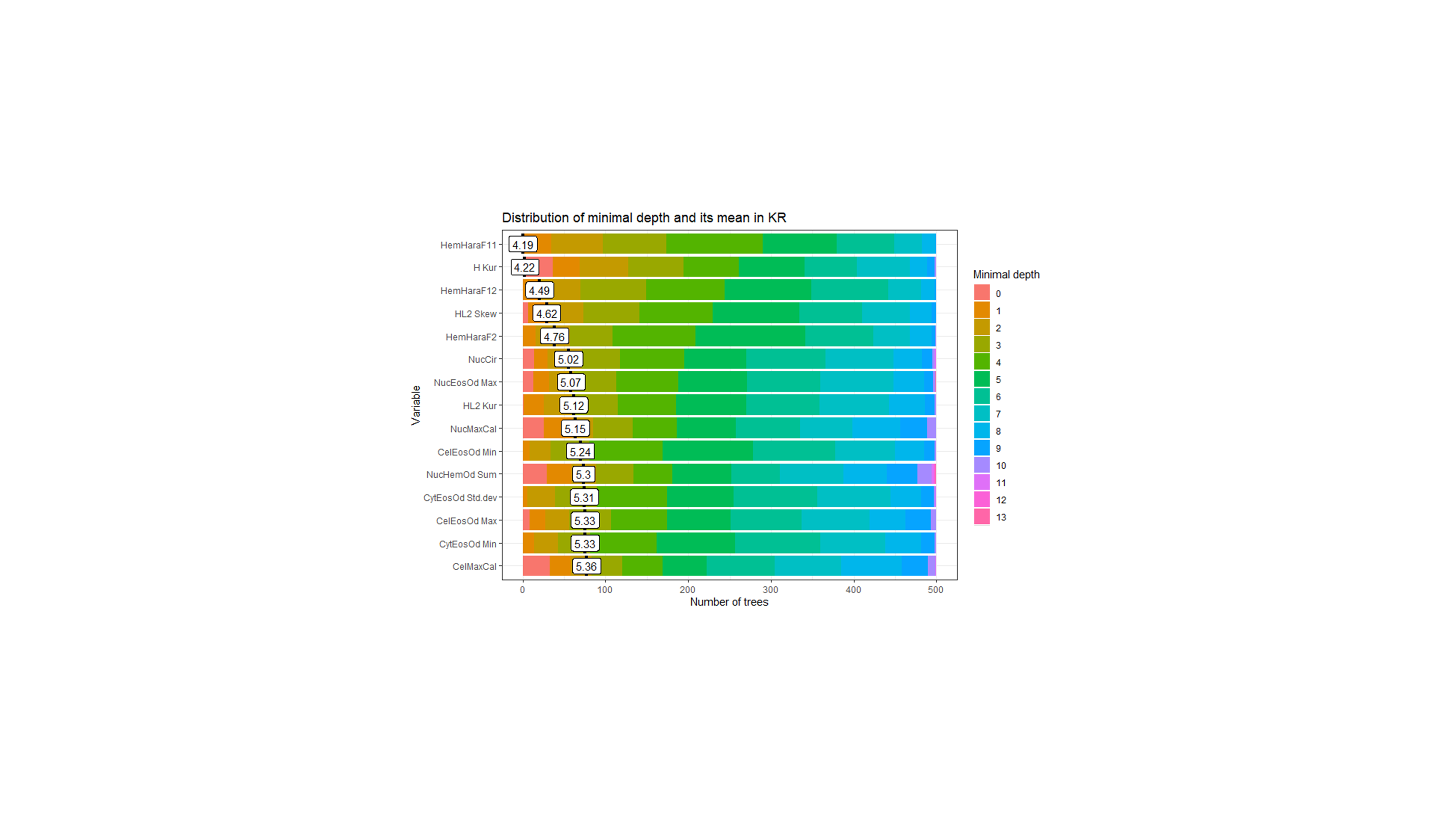

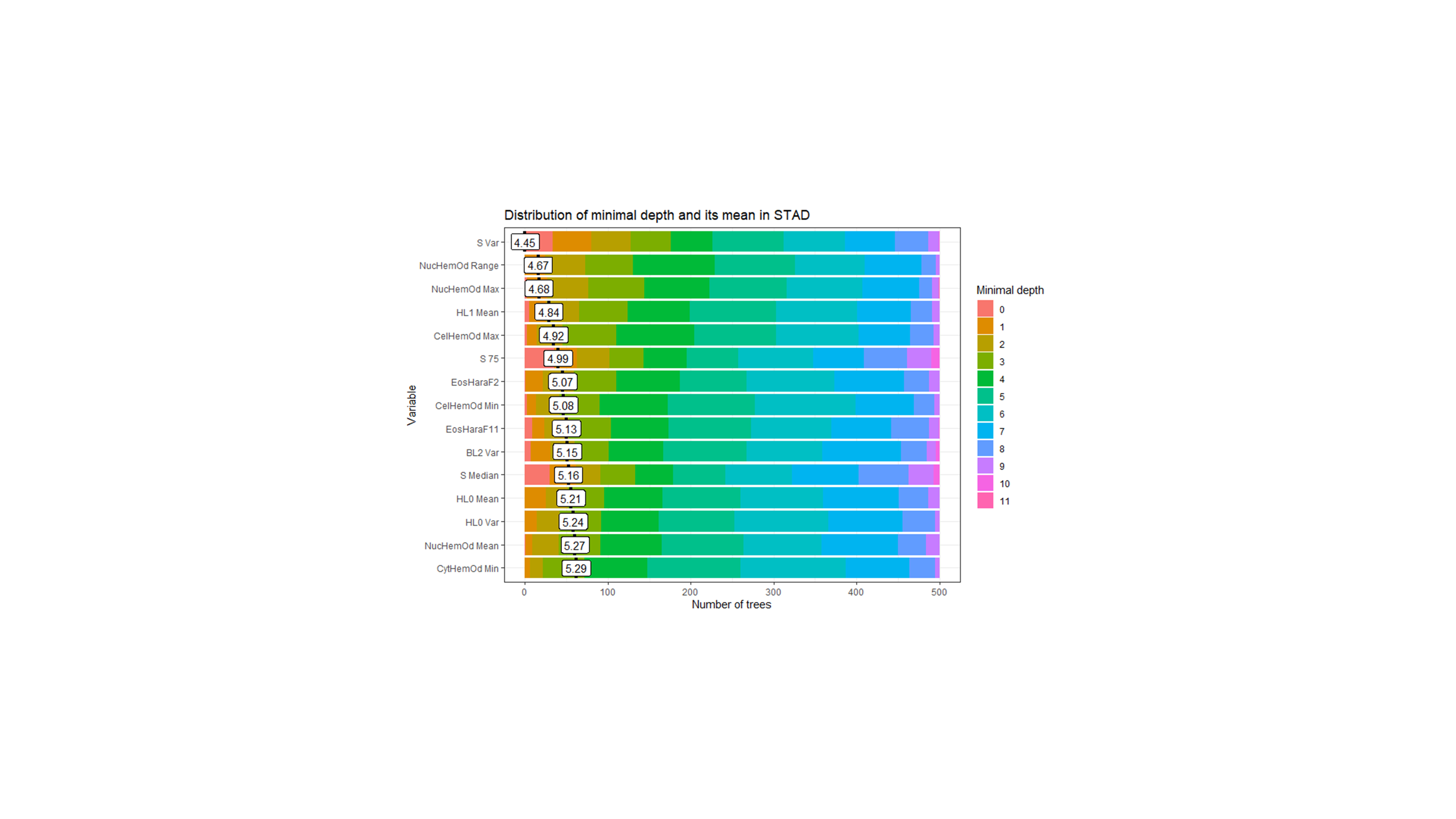

Fig. S4. The distribution of minimal depth for the top ten variables among the trees of random forest in each dataset. The scale of the x-axis goes from zero to the maximum number of trees in which any feature is used for splitting. The length of a bar of the same color counts the number of trees with the corresponding minimal depth. The mean of the distribution is marked by a vertical bar with a value label on it.

Table S1. The abbreviations table.

| **Abbreviations** | **Meanings** |
| --- | --- |
| H | Hue space in HSV color space |
| S | Saturation space in HSV color space |
| V | Value space in HSV color space |
| R | Red space in RGB color space |
| G | Green space in RGB color space |
| B | Blue space in RGB color space |
| HL0 | Hue space in local level 0 from GMM model |
| HL1 | Hue space in local level 1 from GMM model |
| HL2 | Hue space in local level 2 from GMM model |
| SL0 | Saturation space in local level 0 from GMM model |
| SL1 | Saturation space in local level 1 from GMM model |
| SL2 | Saturation space in local level 2 from GMM model |
| VL0 | Value space in local level 0 from GMM model |
| VL1 | Value space in local level 1 from GMM model |
| VL2 | Value space in local level 2 from GMM model |
| RL0 | Red space in local level 0 from GMM model |
| RL1 | Red space in local level 1 from GMM model |
| RL2 | Red space in local level 2 from GMM model |
| GL0 | Green space in local level 0 from GMM model |
| GL1 | Green space in local level 1 from GMM model |
| GL2 | Green space in local level 2 from GMM model |
| BL0 | Blue space in local level 0 from GMM model |
| BL1 | Blue space in local level 1 from GMM model |
| BL2 | Blue space in local level 2 from GMM model |
| Kur | Kurtosis |
| Skew | Skewness |
| Var | Variance |
| Max | Maximum |
| Min | Minimum |
| 25 | The 25% quantile |
| 75 | The 75% quantile |
| Nuc | Nucleus |
| Eos | Eosin part in H&E stain |
| Hem | Hematoxylin part in H&E stain |
| Hara | Haralick features |
| Cyt | Cytoplasm |
| Cir | Circularity |
| Od | Optical Density |
| Std.dev | Standard Deviation |
| Cal | Caliper |
| Ecc | Eccentricity |
| Per | Perimeter |
| Diff | Differentiation |
| F+number | The specific statistics from the Haralick Features |
| Cen | Centroid |
| Cel | Cell |
| Px | Pixel |
| Are | Area |
| X | The horizontal axis |
| Y | The vertical axis |
| Div | Divide |
| DiffLevel | The differentiation level of the tumor tissue |

Table S2. The features’ names sorted by the importance score in each dataset in Fig. 3.a, Fig. S1.a, and Fig. S2.a. The features in the table correspond to the bar in each Figure from above to below.

| **KR** | **DX** | **STAD** |
| --- | --- | --- |
| HL2 Skew | S Skew | S 75 |
| HemHaraF12 | RL0 Skew | NucHemOd Range |
| H Kur | RL0 Var | S Mean |
| HemHaraF11 | CelMaxCal | S Median |
| HemHaraF2 | HemHaraF11 | NucHemOd Max |
| HL2 Kur | B Skew | S 25 |
| CelMaxCal | CelPer | CelHemOd Max |
| NucPer | CelHemOd Max | CytHemOd Min |
| NucAre | NucHemOd Max | CelHemOd Min |
| NucMaxCal | G Skew | HL0 Var |
| NucHemOd Sum | HemHaraF12 | HL1 Mean |
| NucEosOd Max | TumorNum | SL2 Var |
| NucCir | NucMaxCal | EosHaraF2 |
| CytHemOd Max | HemHaraF2 | CelEosOd Max |
| H Skew | NucHemOd Range | EosHaraF11 |
| CelPer | BL2 Var | TumorNum |
| NucEosOd Range | CelAre | BL2 Var |
| CytEosOd Min | HemHaraF4 | HL0 Mean |
| ImmuneNum | EosHaraF2 | NucHemOd Mean |
| CytEosOd Std.dev | HemHaraF8 | NucHemOd Min |
| NucEosOd Std.dev | S Median | EosHaraF12 |
| CelEosOd Min | S Kur | NucEosOd Min |
| NucHemOd Range | EosHaraF11 | CytEosOd Std.dev |
| HL2 Mean | NucPer | H 25 |
| CelAre | NucCir | H Median |
| CelEosOd Max | CytEosOd Std.dev | S Skew |
| HL0 Mean | ImmuneNum | CelEosOd Min |
| CelHemOd Min | CelMinCal | EosHaraF7 |
| CytHemOd Min | BL1 Var | RL0 Var |
| HL1 Kur | HemHaraF0 | HemHaraF11 |
| ImmuneNum Div H 25 | NucHemOd Std.dev | H Mean |
| EosHaraF10 | HemHaraF9 | CytHemOd Mean |
| VL0 Kur | CelEosOd Min | H 75 |
| EosHaraF1 | CelHemOd Min | HL0 Kur |
| HemHaraF1 | RL0 Mean | NucEosOd Max |
| NucMinCal | HemHaraF6 | CytEosOd Min |
| HL1 Var | CytEosOd Min | NucEosOd Range |
| CelHemOd Std.dev | S 75 | B 75 |
| HemHaraF0 | CytHemOd Max | VL0 Var |
| EosHaraF9 | V Skew | HemHaraF2 |
| CytHemOd Mean | NucEcc | NucHemOd Std.dev |
| SL0 Kur | CytHemOd Min | HemHaraF0 |
| SL2 Var | GL2 Var | ImmuneNum |
| NucHemOd Max | S Mean | H Kur |
| CelHemOd Max | NucEosOd Min | BL0 Mean |
| RL2 Kur | NucCelArea Ratio | HemHaraF12 |
| EosHaraF4 | GL1 Var | EosHaraF9 |
| EosHaraF2 | CytHemOd Std.dev | VL1 Kur |
| R 75 | NucHemOd Sum | HL0 Skew |
| RL2 Var | GL0 Mean | SL0 Var |
| EosHaraF11 | S 25 | S Kur |
| TumorNum | H Skew | RL0 Skew |
| CelMinCal | HemHaraF10 | NucHemOd Sum |
| S Kur | HemHaraF1 | EosHaraF10 |
| NucHemOd Min | H Kur | B Mean |
| S Skew | H Median | B Skew |
| CytHemOd Std.dev | EosHaraF10 | GL1 Var |
| CelHemOd Mean | H 75 | VL2 Var |
| H 25 | EosHaraF12 | HemHaraF1 |
| HL2 Var | EosHaraF9 | SL2 Skew |
| R Mean | EosHaraF7 | EosHaraF6 |
| H 75 | H Mean | CelHemOd Mean |
| EosHaraF12 | GL0 Var | SL2 Kur |
| VL2 Var | H 25 | HemHaraF6 |
| NucHemOd Std.dev | EosHaraF1 | RL0 Mean |
| EosHaraF0 | GL0 Skew | VL0 Mean |
| HL1 Mean | NucAre | EosHaraF4 |
| H Mean | EosHaraF4 | HemHaraF5 |
| NucHemOd Mean | CelHemOd Std.dev | CytHemOd Max |
| H Median | HemHaraF7 | RL0 Kur |
| HemHaraF6 | NucEosOd Std.dev | G Kur Div H 25 |
| NucEosOd Min | HemHaraF3 | G 75 |
| HemHaraF8 | NucMinCal | V Skew |
| R 25 | NucEosOd Max | B Median |
| R Kur | CelEosOd Max | HemHaraF10 |
| HemHaraF10 | BL0 Var | EosHaraF1 |
| RL0 Mean | RL2 Skew | G Skew |
| VL0 Mean | BL0 Mean | NucEosOd Std.dev |
| EosHaraF7 | RL1 Skew | SL1 Var |
| HemHaraF3 | NucHemOd Min | CelAre |
| EosHaraF8 | NucEosOd Mean | NucCelArea Ratio |
| RL0 Kur | NucHemOd Mean | HemHaraF4 |
| SL0 Var | R Skew | CelMaxCal |
| SL1 Skew | RL2 Var | G Mean |
| HemHaraF7 | NucEosOd Range | CelMinCal |
| HemHaraF4 | EosHaraF0 | V Kur |
| RL2 Skew | R 75 | RL1 Kur |
| RL1 Var | CytHemOd Mean | CelPer |
| VL0 Skew | RL2 Mean | HemHaraF7 |
| HL0 Var | R Mean | B 25 |
| EosHaraF3 | RL1 Mean | CytHemOd Std.dev |
| NucEosOd Mean | CytEosOd Max | CytEosOd Max |
| HemHaraF9 | RL1 Var | HL2 Var |
| S 75 | R 25 | BL1 Var |
| HemHaraF5 | BL1 Mean | VL0 Skew |
| CelEosOd Std.dev | B Kur | SL2 Mean |
| EosHaraF5 | R Median | SL0 Skew |
| GL2 Kur | EosHaraF5 | GL0 Skew |
| RL1 Skew | ImmuneNum Div H 25 | G Median |
| CytEosOd Max | CytEosOd Mean | CelCir |
| SL0 Mean | BL1 Skew | BL2 Kur |
| SL2 Mean | CelEosOd Mean | GL2 Kur |
| VL2 Mean | BL0 Skew | RL2 Kur |
| SL1 Mean | R Kur | R 75 |
| BL2 Skew | GL1 Skew | RL2 Skew |
| GL0 Var | G Kur Div H 25 | GL0 Mean |
| GL2 Var | CelHemOd Mean | BL2 Mean |
| GL0 Mean | G Kur | VL1 Var |
| BL2 Kur | GL2 Skew | RL1 Mean |
| BL0 Skew | EosHaraF8 | BL0 Skew |
| CelCir | CelEcc | ImmuneNum Div H 25 |
| GL0 Skew | NucEosOd Sum | CelEosOd Std.dev |
| BL0 Mean | EosHaraF6 | EosHaraF3 |
| R Skew | HemHaraF5 | GL2 Mean |
| B Kur | GL1 Mean | RL1 Var |
| BL2 Mean | CelCir | R Median |
| G Kur | BL2 Skew | VL2 Kur |
| GL2 Skew | GL2 Mean | CelEosOd Mean |
| V Skew | B Mean | NucMinCal |
| BL1 Mean | BL2 Mean | R Mean |
| G Median | V Kur | G Kur |
| CenYPx | B 75 | EosHaraF5 |
| G 75 | EosHaraF3 | CytEosOd Mean |
| CenXPx | G 75 | GL1 Kur |
| BL1 Var | V 75 | VL1 Mean |
| G Mean | G Mean | NucPer |
| GL1 Var | G Median | NucMaxCal |
| GL2 Mean | CelEosOd Std.dev | BL0 Var |
| GL1 Mean | V Median | GL1 Mean |
| CelEcc | V Mean | V 25 |
| B Median | B 25 | V 75 |
| V 75 | V 25 | V Median |
| V Median | B Median | V Mean |
| V Mean | G 25 | CenYPx |
| V 25 | CenXPx | CenXPx |
| BL0 Var | CenYPx | CelEcc |
| DiffLevel | DiffLevel | DiffLevel |

Table S3. The features’ names in interaction figures in each dataset in Fig. 3.c, Fig. S1.c, and Fig. S2.c. Each feature pair in the table corresponds to the color blocks in pair separated by a cross in interaction figures from above to below in Figure3, S3 and S4.

| **KR** | | **DX** | | **STAD** | |
| --- | --- | --- | --- | --- | --- |
| **root variable** | **variable** | **root**  **variable** | **variable** | **root**  **variable** | **variable** |
| HL2 Skew | RL2 Kur | S Skew | ImmuneNum | NucHemOd Range | ImmuneNum |
| HL2 Skew | BL1 Kur | RL0 Skew | H 25 | NucHemOd Range | HL2 Var |
| HL2 Skew | HemHaraF12 | S Skew | HemHaraF11 | NucHemOd Range | HL1 Mean |
| HL2 Skew | HL0 Skew | S Skew | RL2 Skew | NucHemOd Range | CelEosOd Max |
| HL2 Skew | H Kur | S Skew | RL1 Skew | NucHemOd Range | H Median |
| HL2 Skew | CelMinCal | S Skew | NucEcc | NucHemOd Range | CelEosOd Min |
| HL2 Skew | HL0 Mean | RL0 Skew | RL0 Var | NucHemOd Range | CelAre |
| HL2 Skew | RL0 Kur | S Skew | GL1 Skew | NucHemOd Range | EosHaraF2 |
| HL2 Skew | HL2 Skew | S Skew | GL0 Mean | NucHemOd Range | HL0 Skew |
| HL2 Skew | ImmuneNum | S Skew | ImmuneNum | NucHemOd Range | CytHemOd Max |
| HL2 Skew | CytHemOd Min | S Skew | EosHaraF2 | NucHemOd Range | NucEosOd Min |
| H Kur | HL2 Skew | S Skew | HemHaraF12 | NucHemOd Range | S Skew |
| HL2 Skew | CelHemOd Max | S Skew | NucHemOd Max | NucHemOd Range | EosHaraF11 |
| HL2 Skew | HL2 Kur | S Skew | RL0 Var | NucHemOd Range | HL2 Skew |
| HL2 Skew | CytEosOd Min | S Skew | RL0 Skew | NucHemOd Range | HL0 Mean |

Table S4. The p-value of each feature in three datasets, respectively, under the Wilcoxon rank sum test. Note that, when the p-value is so small that it exceeds the double-precision floating-point range, it will be 0.

| Feature | P-Value DX | P-Value KR | P-Value STAD |
| --- | --- | --- | --- |
| RL1 Var | 3.59E-267 | 1.00E-52 | 5.94E-34 |
| BL2 Var | 9.80E-256 | 0 | 0 |
| RL2 Var | 0.601107519 | 4.36E-278 | 0.157346181 |
| BL0 Skew | 1.90E-54 | 2.56E-83 | 0 |
| GL0 Var | 9.19E-126 | 2.06E-82 | 7.70E-76 |
| BL1 Var | 0 | 1.34E-35 | 0 |
| BL2 Skew | 0.1472704 | 4.63E-36 | 4.72E-13 |
| GL0 Mean | 5.62E-104 | 0 | 4.15E-296 |
| RL2 Skew | 7.14E-22 | 0 | 3.66E-136 |
| GL1 Var | 8.58E-297 | 2.54E-170 | 0 |
| GL0 Skew | 9.06E-127 | 2.52E-184 | 0 |
| GL1 Mean | 0.350188506 | 0 | 1.57E-90 |
| RL1 Mean | 5.01E-62 | 0 | 6.81E-56 |
| RL0 Var | 8.71E-35 | 1.87E-307 | 2.49E-154 |
| GL1 Skew | 0.353690612 | 4.80E-48 | 6.10E-39 |
| GL2 Mean | 0.044152821 | 3.34E-05 | 0 |
| BL0 Var | 1.95E-133 | 4.06E-115 | 2.80E-176 |
| RL1 Skew | 5.71E-28 | 1.66E-125 | 0.035265802 |
| BL2 Mean | 0.4905311 | 0.001619928 | 1.06E-292 |
| GL2 Skew | 0.000266493 | 1.75E-28 | 2.79E-51 |
| GL2 Var | 3.03E-179 | 0 | 1.82E-266 |
| BL1 Skew | 2.85E-25 | 1.11E-23 | 1.17E-14 |
| BL1 Mean | 0.000233127 | 0 | 4.59E-129 |
| RL0 Skew | 5.21E-277 | 5.71E-29 | 5.39E-258 |
| BL0 Mean | 4.69E-21 | 0 | 0 |
| RL2 Mean | 5.77E-07 | 5.76E-16 | 0 |
| RL0 Mean | 1.10E-217 | 0 | 0.261742479 |
| immunenum | 2.39E-252 | 2.43E-38 | 2.42E-17 |
| spotnum | 1.06E-25 | 1.74E-09 | 4.71E-149 |
| NucEosOd Sum | 0 | 0 | 1.28E-237 |
| EosHaraF1 | 1.45E-29 | 0 | 0 |
| EosHaraF10 | 5.08E-55 | 0 | 0 |
| NucHemOd Std.dev | 0 | 0 | 1.21E-18 |
| NucCir | 0 | 0 | 0 |
| HemHaraF4 | 1.48E-11 | 1.40E-48 | 4.16E-12 |
| HemHaraF8 | 3.28E-186 | 0 | 0.181160256 |
| NucAre | 0 | 0 | 0 |
| CelAre | 0 | 0 | 3.27E-82 |
| EosHaraF7 | 5.97E-37 | 2.42E-223 | 0 |
| CelHemOd Std.dev | 2.15E-05 | 0.314876193 | 8.61E-125 |
| NucEosOd Std_dev | 8.73E-11 | 0 | 0 |
| HemHaraF1 | 2.00E-134 | 0 | 3.53E-14 |
| CelEosOd Std.dev | 1.35E-22 | 8.72E-68 | 0 |
| NucEosOd Range | 3.25E-09 | 0 | 0 |
| EosHaraF8 | 0.431883476 | 9.67E-225 | 0 |
| CytEosOd Std.dev | 5.80E-108 | 2s.64E-78 | 1.96E-07 |
| EosHaraF0 | 7.48E-07 | 2.45E-132 | 0 |
| HemHaraF9 | 0.000164701 | 1.25E-20 | 3.13E-05 |
| CelHemOd Min | 1.59E-59 | 4.45E-103 | 0.200857364 |
| HemHaraF3 | 3.15E-28 | 7.08E-111 | 2.74E-213 |
| CelMaxCal | 0 | 0 | 2.07E-76 |
| NucHemOd Mean | 1.71E-93 | 0 | 3.39E-80 |
| NucMaxCal | 0 | 0 | 6.02E-297 |
| NucHemOd Range | 1.90E-209 | 0 | 0 |
| CelEcc | 0 | 0 | 7.21E-51 |
| HemHaraF0 | 2.87E-45 | 2.55E-207 | 2.78E-14 |
| NucEosOd Mean | 9.89E-19 | 2.31E-22 | 3.81E-06 |
| EosHaraF12 | 1.01E-07 | 1.11E-110 | 0 |
| CelHemOd Mean | 1.56E-189 | 0 | 0.000401154 |
| CelHemOd Max | 5.01E-97 | 8.62E-74 | 0 |
| NucPer | 0 | 0 | 0 |
| CytHemOd Std_dev | 0 | 0 | 7.47E-26 |
| CelEosOd Mean | 3.01E-08 | 1.24E-89 | 0 |
| CelEosOd Min | 4.55E-18 | 1.54E-09 | 1.86E-304 |
| CytEosOd Mean | 1.86E-14 | 1.46E-100 | 0 |
| HemHaraF11 | 9.75E-137 | 1.02E-38 | 3.85E-10 |
| HemHaraF12 | 6.32E-90 | 3.58E-23 | 3.05E-09 |
| EosHaraF2 | 2.71E-174 | 2.01E-23 | 1.89E-273 |
| CenYPx | 0.020626705 | 0.645007505 | 0.441953604 |
| CenXPx | 0.472150747 | 0.315233167 | 0.602055352 |
| NucMinCal | 0 | 0 | 0 |
| CytHemOd Min | 3.15E-59 | 7.09E-102 | 0.034861466 |
| EosHaraF3 | 1.44E-10 | 1.68E-83 | 0 |
| NucEosOd Min | 1.66E-07 | 7.85E-104 | 0 |
| NucHemOd Sum | 0 | 0 | 9.37E-48 |
| HemHaraF7 | 2.07E-163 | 0 | 2.85E-08 |
| NucCelArea Ratio | 3.41E-287 | 0 | 0 |
| EosHaraF11 | 2.32E-28 | 3.86E-11 | 0 |
| CelCir | 0 | 0 | 0 |
| CytEosOd Min | 2.08E-17 | 2.87E-08 | 0 |
| HemHaraF2 | 0.511355632 | 2.90E-158 | 4.57E-165 |
| HemHaraF10 | 2.72E-44 | 4.38E-19 | 0.511991852 |
| NucEosOd Max | 0.17439814 | 0 | 0 |
| CytHemOd Mean | 7.45E-160 | 0 | 4.34E-12 |
| TumorNum | 0 | 0 | 8.92E-05 |
| NucHemOd Min | 1.42E-67 | 0 | 1.45E-16 |
| EosHaraF4 | 1.44E-26 | 0 | 0 |
| CelEosOd Max | 0.001632192 | 0 | 0 |
| NucHemOd Max | 4.29E-108 | 3.85E-76 | 0 |
| HemHaraF5 | 2.35E-176 | 0 | 5.99E-05 |
| EosHaraF6 | 0.005079665 | 2.15E-187 | 0 |
| CytHemOd Max | 0 | 0 | 6.21E-80 |
| NucEcc | 0 | 2.80E-282 | 5.84E-12 |
| EosHaraF5 | 5.64E-08 | 3.80E-76 | 0 |
| CelPer | 0 | 0 | 1.29E-79 |
| CelMinCal | 0 | 0 | 5.78E-55 |
| HemHaraF6 | 6.67E-05 | 7.86E-47 | 5.69E-274 |
| CytEosOd Max | 0.59124779 | 1.63E-58 | 9.46E-269 |
| EosHaraF9 | 2.89E-43 | 0 | 0 |
| H Range | 3.93E-10 | 3.60E-289 | 2.66E-13 |
| R Median | 0.021810527 | 0 | 3.52E-231 |
| R Kur | 2.07E-55 | 0 | 6.49E-53 |
| S Skew | 0 | 0 | 0 |
| R 75 | 0.569351629 | 0 | 3.57E-223 |
| G 25 | 0.006391458 | 0 | 5.13E-164 |
| S Median | 1.51E-120 | 8.88E-293 | 0 |
| B Range | 2.42E-280 | 0 | 0 |
| B 75 | 2.77E-66 | 0 | 0 |
| S Kur | 5.57E-09 | 0.0040273 | 0.01395365 |
| G Var | 2.86E-29 | 0 | 0 |
| V Mean | 4.57E-07 | 0 | 1.05E-106 |
| R 25 | 6.19E-36 | 0 | 8.25E-87 |
| G Skew | 1.47E-179 | 0 | 0 |
| H Median | 2.80E-249 | 0 | 6.97E-236 |
| H 25 | 0 | 0 | 8.75E-281 |
| H Mean | 4.53E-283 | 0 | 5.75E-198 |
| V 25 | 5.37E-19 | 0 | 2.25E-115 |
| V 75 | 0.696136204 | 0 | 3.46E-261 |
| V Range | 7.50E-241 | 2.13E-78 | 7.63E-282 |
| R Var | 7.72E-53 | 2.35E-66 | 0 |
| B Median | 1.53E-62 | 0 | 0 |
| H Skew | 1.87E-273 | 0 | 6.63E-105 |
| S 25 | 3.92E-112 | 0 | 0 |
| R Skew | 5.99E-17 | 0 | 0 |
| G Range | 6.53E-85 | 0 | 0 |
| V Kur | 0.431313439 | 0 | 0 |
| R Range | 0.060903193 | 1.88E-44 | 5.18E-143 |
| H Kur | 2.19E-212 | 0 | 2.57E-60 |
| G Mean | 3.16E-14 | 0 | 0 |
| R Mean | 1.16E-13 | 0 | 1.67E-95 |
| V Var | 6.21E-20 | 1.57E-127 | 0 |
| B Var | 1.00E-80 | 6.25E-178 | 0 |
| B 25 | 0.000152581 | 0 | 3.25E-200 |
| V Median | 0.934911866 | 0 | 3.06E-266 |
| S 75 | 5.95E-43 | 1.37E-103 | 1.60E-251 |
| B Mean | 4.68E-24 | 0 | 0 |
| S Range | 5.17E-224 | 0 | 0 |
| G Median | 5.29E-43 | 0 | 0 |
| B Kur | 4.25E-08 | 6.93E-162 | 2.86E-150 |
| S Mean | 6.72E-65 | 2.22E-238 | 0 |
| S Var | 2.11E-80 | 3.27E-205 | 0 |
| G 75 | 3.52E-37 | 0 | 0 |
| V Skew | 2.26E-07 | 0 | 0 |
| B Skew | 9.97E-167 | 0 | 0 |
| G Kur | 1.53E-60 | 5.25E-35 | 2.60E-69 |
| H 75 | 1.07E-182 | 0 | 1.97E-125 |
| H Var | 1.12E-24 | 0.027584937 | 5.98E-09 |
